# Anesthesia Inmutable Registry of Real-time Vital Signs and Waveforms using Blockchain

**DOI:** 10.1101/2021.12.18.21267876

**Authors:** Alejandro Figar Gutiérrez, Jorge A. Martinez Garbino, Valeria Burgos, Taimoore Rajah, Marcelo Risk, Redelico Francisco

**Affiliations:** Hospital Italiano de Buenos Aires; Instituto Universitario del Hospital Italiano de Buenos Aires; Independent author; Instituto de Medicina Traslacional e Ingenieria Biomedica del Hospital Italiano de Buenos Aires

**Keywords:** anesthesia, blockchain, bitcoin, avionics, tendermint

## Abstract

Healthcare has become one of the most important emerging application areas of blockchain technology.[1] Although the use of a cryptographic ledger within Anesthesia Information Management Systems (AIMS) remains uncertain. The need for a truly immutable anesthesia record is yet to be established, given that the current AIMS database systems have reliable audit capabilities. Adoption of AIMS has followed Roger’s 1962 formulation of the theory of diffusion of innovation. Between 2018 and 2020, adoption was expected to be the 84% of U.S. academic anesthesiology departments.[2] Larger anesthesiology groups with large caseloads, urban settings, and government affiliated or academic institutions are more likely to adopt and implement AIMS solutions, due to the substantial amount of financial resources and dedicated staff to support both the implementation and maintenance that are required. As health care dollars become scarcer, this is the most frequently cited constraint in the adoption and implementation of AIMS.[3]

We propose the use of a blockchain database for saving all incoming data from multiparametric monitors at the operating theatre. We present a proof of concept of the use of this technology for electronic anesthesia records even in the absence of an AIMS at site. In this paper we shall discuss its plausibility as well as its feasibility. The Electronic medical records (EMR) in AIMS might contain errors and artifacts that may (or may not) have to be dealt with. Making them immutable is a scary concept. The use of the blockchain for saving raw data directly from medical monitoring equipment and devices in the operating theatre has to be further investigated.

## Introduction

Most anesthesia data is recorded manually using paper forms, which is time consuming, inefficient, and not registered in real time. Data gathered in the form of paper charting is not portable, access to data is limited and insecure. On the other hand, adoption of EMR systems provides increased improvement of legibility, accurate data capture and improved chart completion. Real-time decision support can be implemented, new opportunities for clinical research arise, there is an improvement in data quality, participation in quality outcomes registries becomes a possibility along with the potential to integrate information across the hospital system.[4] Standards for both paper and electronic medical records require audit trails such that changes are documented.[5] Following this principle, AIMS should retain original values in their underlying databases.

Multiple articles have been published and have long discussed the parallels between anesthesiology and aviation. The first is practiced in operating rooms and the other in airplanes, but both environments require preparedness to manage unexpected situations that put lives at risk, sometimes with incomplete information.[6] In 1992, Hedley stated: “Transfer of information about alarms and monitoring technology should serve several purposes. With both operating room and intensive care patients, a continuous recording system analogous to the flight data recorders used in aeronautics must be developed. Such a system should be non-manipulable by the patients or their attendants and should probably be self-erasing after a specified time limit.” [7]

Those “flight recorders” like systems, mentioned by him 26 years ago, are not standard. AIMS has been developed since, but such systems are not widely available and manual anesthesia records are still being used. There are many systems that would efficiently record anesthesia data in a way that makes it almost impossible to alter electronic records because of the audit capabilities of current database systems. Subsequently, AIMS are still flawed when it comes to recording all the physiological signals emanating from the monitors and primarily focus on numerical data only. The current AIMS has limited capability for the acquisition of high-quality vital signs data. There are software applications like VSCapture or Vital Recorder that could overcome these current disadvantages of AIMS and enable them to support research.[8]

Anesthesia record keeping (developed in the 1890s) has remained an arcane and manual process that relies on the provider to capture and record a wide array of data. The anesthesia record is designed to document not only what was done to a patient during the surgical procedure in the operating room but also how the patient responded. One purported impediment to AIMS adoption is the fear of increased liability risk due to automated inclusion of transient or inaccurate observational data.[3] If there were an adverse outcome, an artifact in the record might be difficult to explain. Editing of the physiological data automatically recorded in AIMS is common practice and results in decreased variability of intraoperative data.[5] Incidence of artifacts and wrong extreme data values, varies depending on the vital signs and also between types of surgical procedures. Artifacts in AIMS databases may influence research results. However, such artifacts can be mitigated by filtration methods during data capturing which can prevent artifacts from being stored.[9] Previous investigations have demonstrated that extreme values may be omitted from handwritten records.[10] Artifact filtration method should be known prior to analyzing AIMS data. Filtering using the median value per minute method provides reliable heart rate and oxygen saturation values, but only acceptable reliability for non-invasive blood pressure.[9] The method of artifact filtering can have substantial effects on estimates of hypotension prevalence. The best way to filter electronically recorded intraoperative blood pressure remains unknown.[11] A prospective clinical study at Wilhelmina Children’s Hospital, Netherlands, found that the incidence of artifacts stored in the present pediatric anesthesia practice was low for heart rate and oxygen saturation, whereas noninvasive and invasive blood pressure and end-tidal carbon dioxide had higher artifact incidences.[11]

Despite many prospective trials on awareness during anesthesia, data from surgeries of patients known to have suffered from it, with a few exceptions, is not readily available. To prevent awareness, dosage thresholds should be established requiring data from anesthesia monitors, end-tidal anesthetic agent, total intravenous anesthesia, including a continuous electronic data stream from induction to emergence.[12]

Blockchain technology is a distributed, decentralized ledger that records transactions. Although the idea for blockchain technology originally appeared with Bitcoin, it has wide applications. In the healthcare area, blockchain is being increasingly explored to optimize business processes, lower overhead costs, improve patient outcomes, enhance compliance, early detection of adverse events and enable better use of health care-related data to improve patient care. [13]

Defined as a chain of blocks (figure 1) that are time-stamped and connected in a peer-to-peer network using cryptographic hashes, a block may contain transactions of many users and generally is publicly available to all users of the network. Each node can be a user or even a computer. Additionally, each block contains the hash of the previous block and the transaction data, thus creating a secure and immutable, append-only chain. This chain continuously increases in length as each new block is being added to its end. Some of the features that characterize blockchain technology are immutability, transparency, auditability, data encryption and operational resilience. Blockchain harnesses the power of encryption to assert its own immutability and becomes a safer option than any physical database.

**Figure 1.**
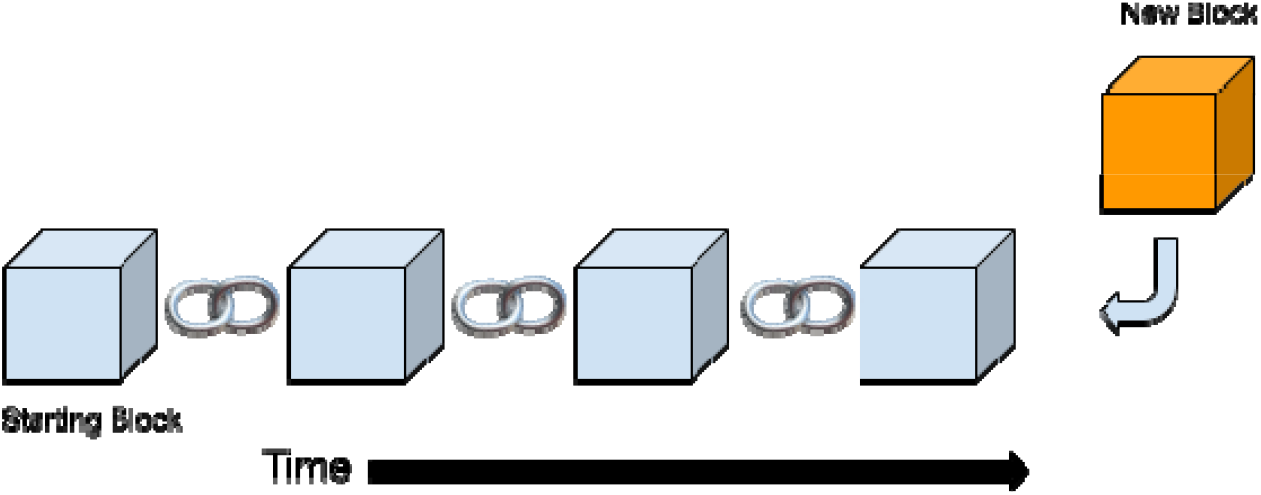
The blockchain technology: a chain of blocks that are time-stamped and connected in a peer-to-peer network using cryptographic hashes.

Healthcare has become one of the most important emerging application areas of blockchain technology.[1] Some uses of blockchain have been targeted to: (a) prescription tracking to detect opioid overdose and over prescription, (b) data sharing to integrate telemedicine with traditional care, (c) sharing cancer data with providers using patient authorized access for targeted therapy, (d) cancer registry sharing to aggregate observations in cancer cases, (e) management of patient digital identity for better patient record matching, (f) personal health records for accessing and controlling complete health history and (g) adjudication automation of health insurance claims to surface error and fraud.[14] Blockchain places the patient at the center of the healthcare ecosystem more than any other systems.[15] One relevant feature of the blockchain technology that makes it very useful in healthcare is its decentralized management because the organization of digital assets and operations produced by various institutions can be performed without a hub.

At present, most of the studies regarding the application of blockchain in medicine are in the conceptual or architectural design phase. Only a few studies report real-world demonstration and evaluation of such technology.[16] Research is mainly focused on integration, integrity and access control of health records and related patient data. However, other diverse and interesting applications are emerging which are addressing medical research, clinical trials, medicine supply chain and medical insurance.[17]

To initiate the design and building of a real-world healthcare blockchain project, one of the critical steps is to select the most suitable underlying blockchain platform. The three most popular platforms currently available in the market are: Ethereum, Hyperledger, and MultiChain. [18]

## Material and Methods

BigchainDB is a scalable blockchain database.[19] It is designed to merge the best of two worlds: the “traditional” distributed database world and the “traditional” blockchain world. The Interplanetary Database Foundation (IPDB) promotes science and research on the operation of scalable blockchain databases, the governance of decentralized systems, and the applications of blockchain technology.[20] It offers a testing site exposing a RESTful application interface that allows us to save the above mentioned data: https://test.ipdb.io/, operated by IPDB. Due to The General Data Protection Regulation (GDPR) reasons, in the European Union and Economic Area, it will be reset every day at 4 am CET. We have installed one instance at http://test-bigchaindb.for-our.info:9984/ for this project, but the IPDB Foundation testnet can also be used. BigchainDB nodes communicate with each other using *Tendermint* wire protocols (figure 2). [21,22]

**Figure 2:**
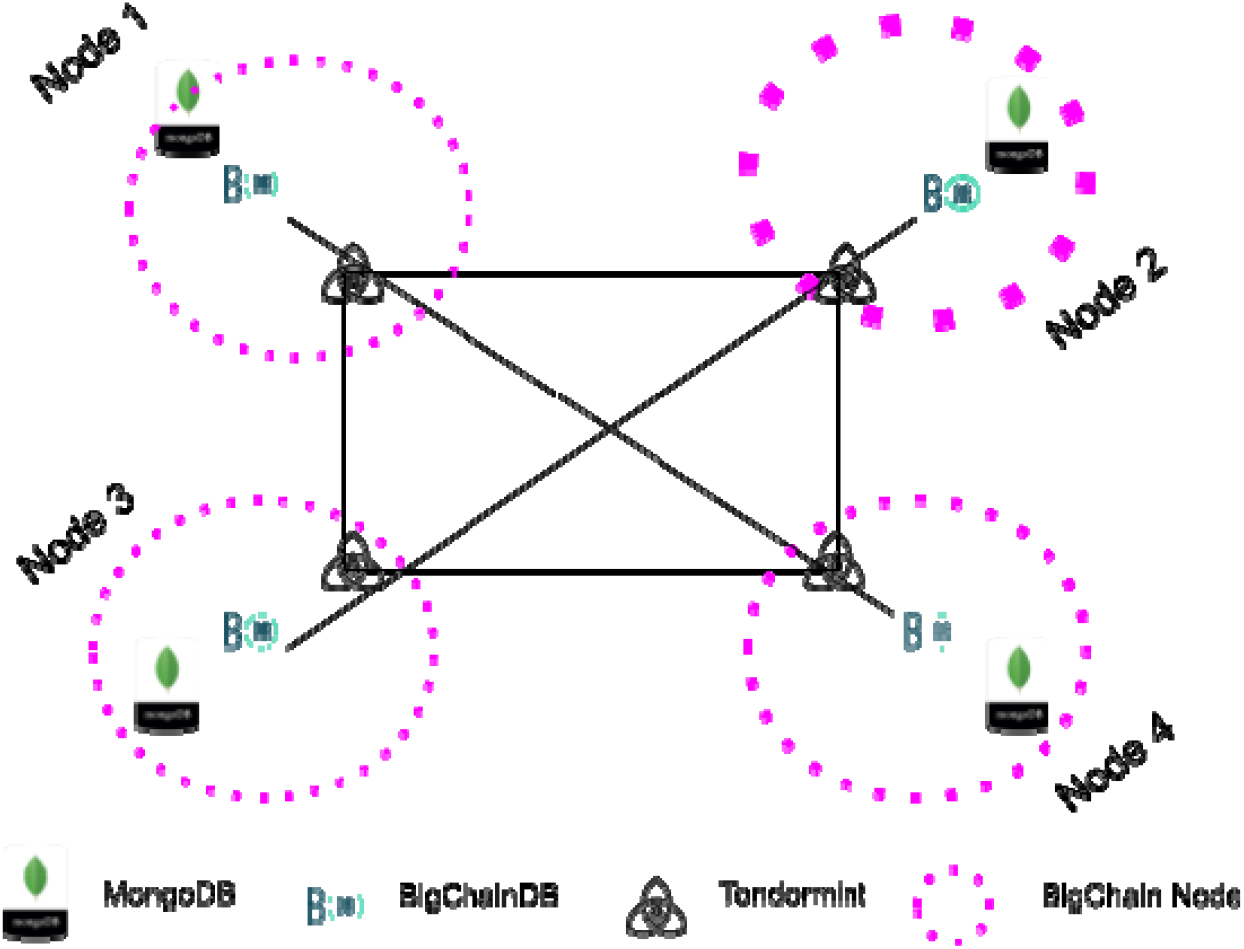
BigChainDB architecture.

Until recently, building a blockchain required building all three layers (*Networking, Consensus*, and *Application*) from the ground up. Tendermint Byzantine Fault Tolerant (BFT), created by Jae Kwon in 2014 collaborating with Ethan Buchman, a solution that packages the *networking* and *consensus* layers of a blockchain into a generic engine, allowing developers to focus on the *application* development aspect as opposed to dealing with the complex underlying protocol.

Bitcoin is noted for its ability to withstand attacks and malicious behaviors by any of the participants.[23] Traditionally, consensus protocols tolerant of malicious behavior were known as BFT consensus protocols. The term Byzantine was used due to the similarity of the problem that faced the generals of the Byzantine army attempting to coordinate themselves to attack Rome using only messengers, where one of the generals may be a traitor.[24]

Google colab or “the Colaboratory” is a free cloud service hosted by Google to encourage machine learning and artificial intelligence research, where often a barrier to learning and success is the requirement of tremendous computational power.[25] Collaboration feature (works with a team just like Google Docs): Google Colab allows developers to use and share Jupyter notebook among each other without having to download, install, or run anything other than a browser. The Jupyter Notebook project is an open-source web application that allows you to create and share documents that contain live code, algorithms, equations, visualizations, and narrative text. Uses include data cleaning and transformation, numerical simulation, statistical modeling, data visualization, machine learning, and much more.[26]

We used VSCapture, a free open source software to get one minute of pleth waveform and DII ECG lead from a healthy volunteer using a multi-parametric monitor.[27,28] This software has many capabilities, it is able to output data into many formats like csv (comma separated) files and it is also able to output data streams to a RESTful API (an application program interface that uses HTTP requests to GET, PUT, POST and DELETE data) using the POST method. We developed one serverless lambda amazon web services nodejs application, which exposes an API to receive output from VSCapture. This lambda function sends data to an Amazon Simple Notification Service (SNS). Each arriving message triggers a second lambda function that formats the received messages adding metadata to them, as required by the bigchainDB blockchain database. Incoming messages are escaped, stringified and wrapped in JSON format, then it is set as an ‘asset’ owned by a patient named ‘alice’. We update metadata of a patient’s asset in the blockchain database as a new record or event arrives. Metadata consists of patient name, monitor unique id, location, and operating room (figure 3). This architecture is for demonstration purposes only and it is not recommended for a production environment. NodeJs code for both lambda functions is available at Github public repositories. [29,30]

**Figure 3:**
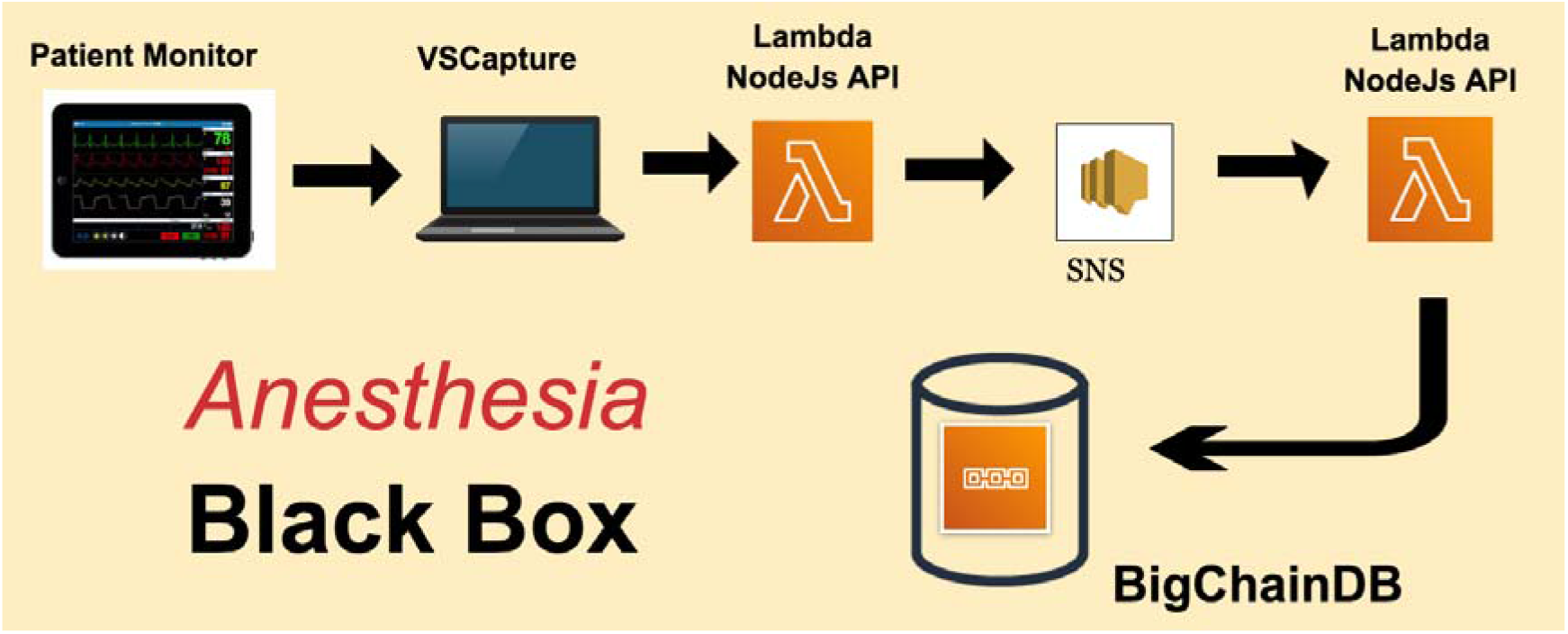
Data workflow.

First lambda function entry point is at: https://r2azbfz0j5.execute-api.us-east-1.amazonaws.com/dev/api/v1/

At the moment of this writing VSCapture software is not able to POST requests to a secure web server (HTTPS protocol). For demonstration purposes we provide a VSCapture executable modified to enable it to use HTTPS protocol (required by lambda functions) at: https://drive.google.com/file/d/1SW7lc6vUhYb9gbh0a8VRJafBSgD0SEvB/view?usp=sharing

Our collaboration with VSCapture project on this matter can be read at: https://sourceforge.net/p/vscapture/discussion/general/thread/f1412ace2c/

On the other hand, we provide an entry point that is able to receive unsecured POST requests, that might work with unmodified version of VSCapture at: http://test-lambda.for-our.info:50000/api/v1/

A python Jupyter notebook [31] within a google colab environment is publicly available to retrieve sent information, querying by different criteria: metadata or physiological Id. This might not be user friendly, and some technical knowledge might be necessary to run this code: https://colab.research.google.com/drive/1YWIppzi-lL5rVk0bOvnNAJEWJB0P5zOT

We have made all necessary code available in a public GitHub repository, in order to let others, replicate this experience. We also provide a Jupyter notebook with sample data for saving it on BigchainDB test site, as we know that test site is sometimes shut down and started a new, empty one, just in case you do not want to replicate the whole experiment, at: https://colab.research.google.com/drive/1qUpGdmq1GLXKrCKXLDPYbQLtSmPKmqKP

## Results and Discussion

AIMS are not widely available, although adoption rate is on the rise worldwide. Electronic devices that have real time data export capabilities are almost standard equipment at the operating theatre. The Internet of Things (IoT: the collection and exchange of data between inter-connected physical devices via internet protocol) turns data into action: all data should be captured, emitted, transported, stored (in the cloud). Even artifacts and noise could be useful for machine learning algorithms. Every data point is valuable and should be kept for future analysis. Physiological data streams should be treated the same way. Although there are many AIMS already doing this, there are multiple vendors, with different data export protocols. We aim to present the concept that saving raw physiological data to a blockchain database is feasible, and it could guarantee data immutability and encryption to preserve patient privacy and other characteristics inherited from the hyperledger technology (blockchain). For this purpose, we present a proof of concept, to demonstrate that it is technically possible to create this functionality.

Being able to retrieve data recorded from all devices connected to a patient, along with data from environmental sensors, during anesthesia from a distributed, decentralized blockchain database, could be the foundation of the future “Operating Room Blackbox”, just like a flight recorder in an airplane. There are blockchain managed solutions that allow us to focus on the application. For example, BigchainDB has high throughput: with 32 nodes, the write throughput was just over 1 million writes per second (i.e., 1,000 blocks per second, with 1,000 transactions per block), high capacity, rich permissioning, query capabilities and low latency. [21] Thus It might be a good choice for storing data streams using blockchain while preserving query capabilities of a NOSQL database.

The actual need of a blockchain database for recording data streams from multiparametric monitors at the operating theatre is yet to be demonstrated. It seems actual systems are up to the task and are not easy to be tampered with. Immutability of anesthesia records seems to be controversial. From a medico-legal standpoint, there is no evidence that routine editing of data is superior to defending a claim that artifacts and transient readings are frequent occurrences that usually lack clinical significance.[5]

Raw physiologic data might be used for feature extraction to feed data to machine learning models developed for different purposes, like algorithms designed for predicting hypotension at the operating theater. Thus, moving from “detection and reaction” to “proactive treatment” of adverse events like an impending hypotension, could improve health outcomes on surgical patients.[32] There are many statistical methods for dealing with outliers in time series data during the data wrangling and the feature extraction phase for building machine learning models, that could also be considered for filtering raw physiological data, before saving to AIMS databases.[33] Any use of this data for research purposes has to take into account AIMS data filtering policies that could influence research results. Dealing with artifactual data is a concern among authors. The use methods learned from predictive analytics in recent years for dealing with artifacts should be further explored, as long as the role of neural networks.

This could be an adequate solution for sites without AIMS, that could fill in the gap until one of such systems is implemented. Operating room devices should be considered as IoT ones, whose raw data should be emitted to a queue ending up in an immutable database. This might be a scary concept, because once the data is in the blockchain, it is immortal. It can be updated to reflect changes, but all entries are new, and are also indelible events. If you have an AIMS doing automatic data filtering, this blockchain immutable database could provide raw data for research and even to audit AIMS databases to improve accuracy of automatic filtering algorithms.

## Data Availability

https://colab.research.google.com/drive/1YWIppzi-lL5rVk0bOvnNAJEWJB0P5zOT

## References

1. Kuo T-T, Kim H-E, Ohno-Machado L. Blockchain distributed ledger technologies for biomedical and health care applications. J Am Med Inform Assoc. 2017;24: 1211–1220. doi:10.1093/jamia/ocx068

2. Stol IS, Ehrenfeld JM, Epstein RH. Technology diffusion of anesthesia information management systems into academic anesthesia departments in the United States. Anesth Analg. 2014;118: 644–650. doi:10.1213/ANE.0000000000000055

3. Trentman TL, Mueller JT, Ruskin KJ, Noble BN, Doyle CA. Adoption of anesthesia information management systems by US anesthesiologists. J Clin Monit Comput. 2011;25: 129–135. doi:10.1007/s10877-011-9289-x

4. Bloomfield EL, Feinglass NG. The anesthesia information management system for electronic documentation: what are we waiting for? J Anesth. 2008;22: 404–411. doi:10.1007/s00540-008-0643-1

5. Wax DB, Beilin Y, Hossain S, Lin H-M, Reich DL. Manual editing of automatically recorded data in an anesthesia information management system. Anesthesiology. 2008;109: 811–815. doi:10.1097/ALN.0b013e3181895f70

6. Eltorai AS. Lessons from the sky: an aviation-based framework for maximizing the delivery of quality anesthetic care. J Anesth. 2018;32: 263–268. doi:10.1007/s00540-018-2467-y

7. Hedley-Whyte J. Operating Room and Intensive Care Alarms and Information Transfer. ASTM International; 1992. Available: https://market.android.com/details?id=book-gnbT_Q4E7YEC

8. Lee H-C, Jung C-W. Vital Recorder-a free research tool for automatic recording of high-resolution time-synchronised physiological data from multiple anaesthesia devices. Sci Rep. 2018;8: 1527. doi:10.1038/s41598-018-20062-4

9. Kool NP, van Waes JAR, Bijker JB, Peelen LM, van Wolfswinkel L, de Graaff JC, et al. Artifacts in research data obtained from an anesthesia information and management system. Can J Anaesth. 2012;59: 833–841. doi:10.1007/s12630-012-9754-0

10. Lerou JG, Dirksen R, van Daele M, Nijhuis GM, Crul JF. Automated charting of physiological variables in anesthesia: a quantitative comparison of automated versus handwritten anesthesia records. J Clin Monit. 1988;4: 37–47. doi:10.1007/bf01618106

11. Hoorweg A-LJ, Pasma W, van Wolfswinkel L, de Graaff JC. Incidence of Artifacts and Deviating Values in Research Data Obtained from an Anesthesia Information Management System in Children. Anesthesiology. 2018;128: 293–304. doi:10.1097/ALN.0000000000001895

12. Nickalls RWD, Mahajan RP. Awareness and anaesthesia: think dose, think data. Br J Anaesth. 2010;104: 1–2. doi:10.1093/bja/aep360

13. Mackey TK, Kuo T-T, Gummadi B, Clauson KA, Church G, Grishin D, et al. “Fit-for-purpose?” -challenges and opportunities for applications of blockchain technology in the future of healthcare. BMC Med. 2019;17: 68. doi:10.1186/s12916-019-1296-7

14. Hughes F, Morrow MJ. Blockchain and Health Care. Policy Polit Nurs Pract. 2019; 1527154419833570. doi:10.1177/1527154419833570

15. Linn LA, Koo MB. Blockchain For Health Data and Its Potential Use in Health IT and Health Care Related Research. Available: https://www.healthit.gov/sites/default/files/11-74-ablockchainforhealthcare.pdf

16. Mettler M. Blockchain technology in healthcare: The revolution starts here. 2016 IEEE 18th International Conference on e-Health Networking, Applications and Services (Healthcom). 2016. pp. 1–3. doi:10.1109/HealthCom.2016.7749510

17. Drosatos G, Kaldoudi E. Blockchain Applications in the Biomedical Domain: A Scoping Review. Comput Struct Biotechnol J. 2019;17: 229–240. doi:10.1016/j.csbj.2019.01.010

18. Kuo T-T, Zavaleta Rojas H, Ohno-Machado L. Comparison of blockchain platforms: a systematic review and healthcare examples. J Am Med Inform Assoc. 2019. doi:10.1093/jamia/ocy185

19. BigchainDB • • The blockchain database. In: BigchainDB [Internet]. [cited 18 Apr 2019]. Available: https://www.bigchaindb.com/

20. Foundation & governance · IPDB. In: IPDB [Internet]. [cited 30 Jul 2019]. Available: https://ipdb.io/foundation/

21. Buchman E. Tendermint: Byzantine Fault Tolerance in the Age of Blockchains. Taylor G, editor.Master of Applied Science in Engineering Systems and Computing, University of Guelph, Ontario, Canada. 2016. Available: https://atrium.lib.uoguelph.ca/xmlui/bitstream/handle/10214/9769/Buchman_Ethan_201606_MAsc.pdf

22. tendermint. Github; Available: https://github.com/tendermint/tendermint

23. McGinn D, McIlwraith D, Guo Y. Towards open data blockchain analytics: a Bitcoin perspective. R Soc Open Sci. 2018;5: 180298. doi:10.1098/rsos.180298

24. Lamport L, Shostak R, Pease M. The Byzantine generals problem. ACM Trans Program Lang Syst. 1982;4: 382–401. Available: https://people.eecs.berkeley.edu/~luca/cs174/byzantine.pdf

25. Google Colaboratory. [cited 18 Apr 2019]. Available: https://colab.research.google.com/notebooks/welcome.ipynb

26. Project Jupyter. [cited 18 Apr 2019]. Available: https://jupyter.org/

27. Karippacheril JG, Ho TY. Data acquisition from S/5 GE Datex anesthesia monitor using VSCapture: An open source.NET/Mono tool. J Anaesthesiol Clin Pharmacol. 2013;29: 423–424. doi:10.4103/0970-9185.117096

28. VSCaptureMP. Github; Available: https://github.com/xeonfusion/VSCaptureMP

29. Gutierrez AF. anesthesia-blockchain-sns. Github; Available: https://github.com/afigar/anesthesia-blockchain-sns

30. Gutierrez AF. anesthesia-blockchain. Github; Available: https://github.com/afigar/anesthesia-blockchain

31. Gutiérrez AF. Anesthesia Blockchain Inmutable Registry BigchainDB.ipynb. [cited 20 Sep 2019]. Available: https://colab.research.google.com/drive/1YWIppzi-lL5rVk0bOvnNAJEWJB0P5zOT

32. Vistisen ST, Johnson AEW, Scheeren TWL. Predicting vital sign deterioration with artificial intelligence or machine learning. Journal of clinical monitoring and computing. 2019. pp. 949–951. doi:10.1007/s10877-019-00343-7

33. Chen C, Liu L-M. Joint Estimation of Model Parameters and Outlier Effects in Time Series. J Am Stat Assoc. 1993;88: 284–297. doi:10.1080/01621459.1993.10594321

